# Toward Interpretable Schizophrenia Detection from EEG Using Autoencoder and EfficientNet

**DOI:** 10.1101/2025.05.16.25327657

**Authors:** Umesh Kumar Naik Mudavath, Shaik Rafi Ahamed, KongFatt Wong-Lin

## Abstract

Schizophrenia (SZ) is a complex mental disorder that necessitates accurate and timely diagnosis for effective treatment. Traditional methods for SZ classification often struggle to capture transient EEG features, face high computational complexity, and lack interpretability. This study proposes two complementary pipelines, one using a convolutional autoencoder (CAE) combined with an extreme gradient boosting (XGB) classifier. Alternatively, we introduce a unique approach employing spectral scalograms (SS) combined with the EfficientNet (ENB) architecture. The SS, obtained through continuous wavelet transform, reveals temporal and spectral information of EEG signals, aiding in the identification of transient features, aiding in accurate SZ classification. Experimental evaluation on a comprehensive dataset demonstrates the efficacy of our approach, achieving a five-fold mean cross-validation accuracy of 95.3% using CAE with XGB and 97% utilizing SS with the ENB0 model. Grad-CAM was then applied on ENB0 to highlight the time-frequency bands key to each decision, and SHAP on the CAE-XGB pipeline to rank the most essential EEG channels. These complementary views clarify both where and why the models work, paving the way for more transparent clinical EEG analysis.

## Introduction

Schizophrenia (SZ) is a debilitating mental disorder marked by disturbances in thought processes, emotions, and behavior [1]. It typically emerges between ages 16 and 30, often following a first episode of psychosis. Subtle changes in thinking and mood may precede this episode. Early and accurate detection is crucial for timely and appropriate treatments and care, including antipsychotic medications, psychotherapy, and support services [1]. Diagnosis follows DSM-5-TR criteria, which require at least two of the following symptoms: delusions, hallucinations, disorganized speech, grossly disorganized or catatonic behavior, or negative symptoms, present for one month, with overall disturbance lasting at least six months [2].

Despite decades of research, the exact cause remains unclear [3], though structural brain abnormalities are well-documented, including ventricular enlargement, reduced cortical and hippocampal volume, and altered cerebral asymmetries [4]. EEG studies in SZ have shown increased theta and delta waves [5], decreased alpha waves [6], and abnormalities in event-related potentials (ERPs) [7]. However, the disorder’s complexity often leads to delayed or missed diagnoses. Delayed treatment can worsen symptoms and functional decline, even with pharmacological intervention [3]. Childhood factors, such as delayed motor development, speech issues, social anxiety, and low academic performance, are linked to elevated SZ risk in adulthood [8].

EEG has become a valuable tool in identifying SZ biomarkers and analyzing brain electrical activity [5]. It captures neural signals that reveal abnormalities associated with the disorder. By examining patterns, frequencies, and connectivity in EEG data, researchers can detect markers of SZ. Analytical methods include spectral analysis, ERPs, and connectivity measures, often combined with machine learning (ML) to identify distinctive EEG features, such as altered oscillatory rhythms [5, 6], ERP abnormalities [7], and disrupted connectivity [9]. Deep learning (DL) models have also gained traction, leveraging powerful neural networks to learn patterns from EEG signals and detect SZ from healthy controls (HC).

Researchers have explored both hand-crafted features (HCF) and latent feature approaches [10]. Common HCFs include statistical and time-frequency measures like Shannon entropy, dominant frequency, and spike rhythmicity [11]. For example, Yang et al. [12] developed a hybrid ML method combining functional magnetic resonance imaging (fMRI) and single nucleotide polymorphism data, selecting discriminative features and constructing support vector machine (SVM) ensembles, which improved classification accuracy over single-modality approaches. As standard univariate analysis of neuroimaging data has limitations in clinical use, Orru et al. [13] reviewed alternative approaches, particularly SVMs, showing promise in diagnosis, prognosis, and transition prediction. Later, Siuly et al. [14] developed SchizoGoogLeNet, using GoogLeNet-based feature extraction, achieving 98.84% accuracy on EEG data.

Current automatic classification approaches often rely on simplistic learning models that struggle with the inherent complexity of non-stationary EEG data, resulting in limited adaptability and poor generalization. Many techniques fail to capture the multi-resolution nature of EEG signals and, importantly, lack the ability to gain insights from so-called black box models, which are often criticized for their inability to explain how they arrive at a particular decision. However, the narrative has shifted; we can now not only leverage these frameworks but also gain explanations from them. Our approach combines signal decomposition with ML to accurately analyze EEG data in SZ while also offering interpretability. To address these challenges, we propose a two-fold approach. One part leverages the combination of discrete wavelet transform (DWT) with a convolutional autoencoder (CAE) and ensemble models. On the other hand, EfficientNetB0 (ENB0) with spectral scalograms (SS) [15]. Wavelet-based methods, including continuous and discrete wavelet transforms, are useful in capturing both temporal and spectral EEG characteristics [16]. These offer a detailed view of frequency components over time, supporting the identification of SZ-related patterns and serving as potential biomarkers. Fig. 1 depicts the proposed classification framework. The multi-channel EEG data is decomposed into sub-bands via DWT, reducing redundancy and enabling efficient analysis. These sub-bands are fed into a CAE for sparse representation, lowering computational cost and supporting real-time use. Simultaneously, SS generated employing continuous wavelet transform (CWT) preserves rich temporal and spectral information. These are later fed to the ENB0 model, which works based on transfer learning (TL). The ENB model allows deep feature extraction from SS, reducing manual feature engineering [15], and supporting the model in learning hierarchical patterns to distinguish between SZ and HC [17]. It also helps the model generalize to smaller biomedical datasets by leveraging pre-trained knowledge, which reduces reliance on large labelled data, a common limitation in SZ research.

**Fig. 1.**
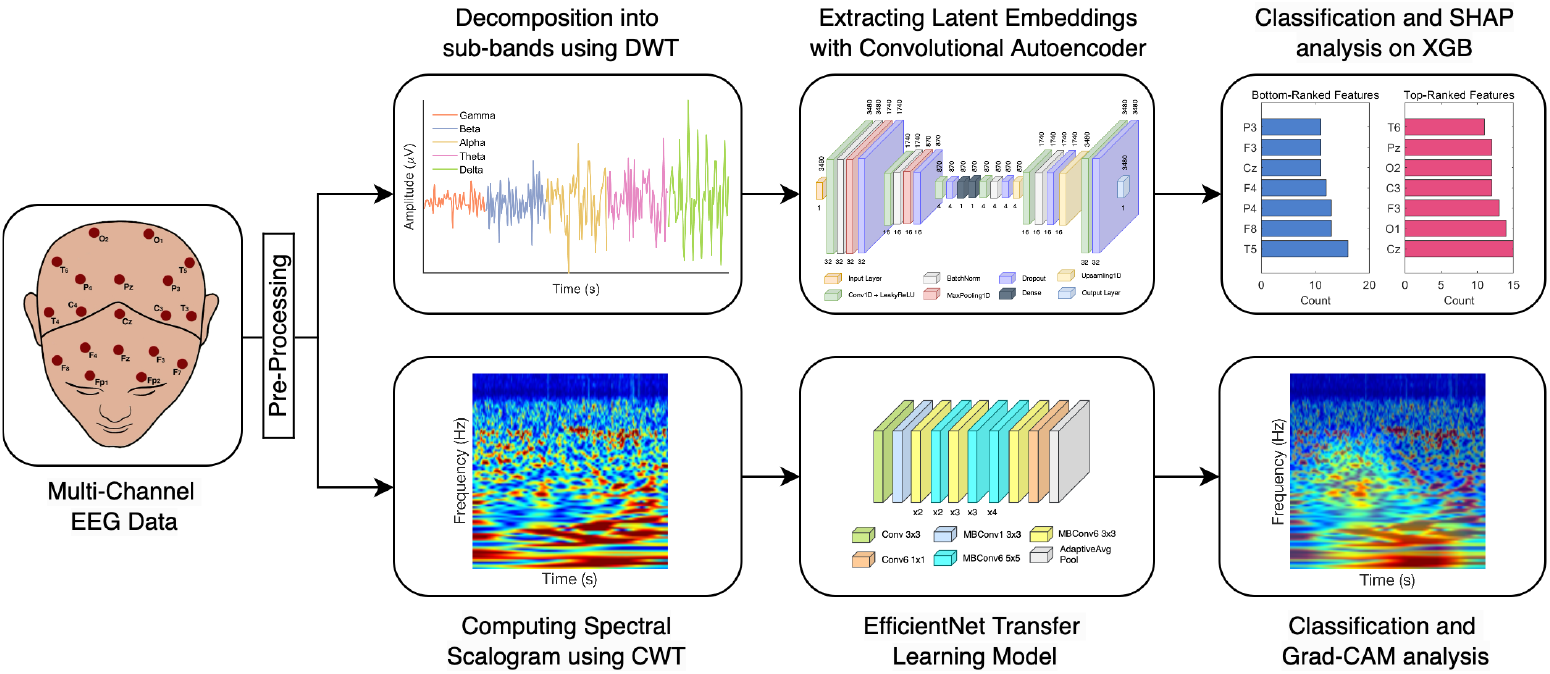
Interpretable EEG-based schizophrenia detection pipeline. Top: DWT-decomposed EEG sub-bands are encoded by a convolutional autoencoder (CAE) to extract latent embeddings, which are classified with extreme gradient boosting (XGB). SHapley Additive exPlanations (SHAP) then highlight the most and least influential features. Bottom: CWT-derived spectral scalograms (SS) are classified by EfficientNetB0 (ENB0), with Gradient-weighted Class Activation Mapping (Grad-CAM) indicating the essential time-frequency segments for each decision. The multichannel EEG head model is adapted from [20].

High accuracy alone is not enough to build trustworthy artificial intelligence (AI) models. Explainable AI (XAI) techniques are, ergo, essential to reveal how a model arrives at its decisions and to foster clinician trust. To address this, Gradient-weighted Class Activation Mapping (Grad-CAM) [18] was integrated into the SS-ENB pipeline. As a visual explanation technique in XAI, Grad-CAM was used to overlay heatmaps on SS images to show which time-frequency patches the network found most relevant, helping clinicians understand the model’s decision-making process. Additionally, we used SHAP [19], another feature attribution method within XAI, to interpret CAE-XGB predictions by ranking the most influential EEG channels contributing to classification. This EEG-based diagnostic approach aims to enhance early detection, capture transient EEG features, improve interpretability, address computational complexity, and contribute to understanding SZ neurobiology. It offers a promising direction for more objective and accessible clinical assessments.

## Methodolgy

### Dataset description

The open dataset [20] used in this study, recorded at 128 Hz from 16 electrodes placed according to the 10-20 channel system (Fig. 1), focuses on adolescents aged from 10 to 14. The first group includes 45 boys (ages from 11 to 14) diagnosed with SZ, and the second group includes 39 healthy boys (ages between 10 to 13). EEG segments were collected from awake adolescents in a relaxed, eyes-closed state. To ensure EEG integrity, none of the subjects had taken medication prior to the study. See the original source [20] for further details.

### Data pre-processing

Initially, the EEG data of all subjects were normalized to eliminate scaling differences and ensure consistency for accurate analysis. The original EEG signal, with a length of 122880 per subject, was reshaped into a 16×7680 matrix. Each of the 16 rows corresponds to one EEG channel, with 7680 samples per channel, as specified in the dataset description [20]. For the CAE approach, EEG signals were decomposed into delta, theta, alpha, beta, and gamma bands [11], and the resulting components were concatenated before being fed into the CAE for training. For the ENB0 model, a CWT filter bank was used to convert the EEG time series into 2D SS images. The filter bank used the ‘bump’ wavelet with 12 voices per octave, a sampling frequency of 128 Hz, and a frequency range of 0.5-50 Hz. These parameters enabled effective analysis of the EEG signals. The resulting SS were used as input to the ENB0 model for classification.

### Wavelets

#### DWT for decomposition

DWT [21] was employed since it offers superior multiresolution capability, enabling signal decomposition into different frequency bands at various resolutions. It also provides better time-frequency localization and is more effective at reducing noise in signals, and is particularly suitable for evaluating non-stationary signals like EEG [11]. By segmenting the EEG signal into diverse frequency bands followed by downsampling, DWT allows for efficient analysis with reduced time complexity (Fig. 1).

#### CWT for Scalogram

CWT [22] was used to transform EEG signals into SS, depicting how signal energy evolves over both time and frequency (Fig. 1). By decomposing the signal with a family of scaled and shifted wavelets, CWT provides adaptive resolution, using wide windows for slow components and narrow windows for fast ones, making it well suited to capture transient features of SZ-related brain activity. This flexibility and superior time-frequency localization help uncover subtle patterns that are not apparent in the raw signal or in fixed-window methods like short-time Fourier transform (STFT) [23].

#### Convolutional autoencoder

The CAE comprises an encoder and decoder network trained to learn compact hierarchical representations of the input EEG data. Specifically, a wavelet-based CAE was employed that uses DWT to pre-process the EEG data. Multivariate features are extracted from the bottleneck layer and used as input to extreme gradient boosting (XGB) [24] for classification (Fig. 1). Overall, the CAE provides a data-driven and efficient approach for SZ classification from EEG, supporting accurate diagnosis and a deeper understanding of this disorder. Mathematically, let 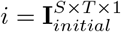 represent the input EEG data, where *T* is the number of time points and *S* is the number of samples. Let E and D represent the Encoder and Decoder of the CAE. The encoder represen-tation is 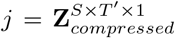, where *T* ^*′*^ is the compressed temporal representation, and the decoder reconstruction is 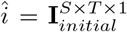. These can be expressed as *j* = *E*(*i*) and *î* = *D*(*j*). The reconstruction error *r*^*wDAE*^ used to evaluate the CAEs’ performance, given by:

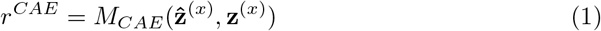

The *M*_*CAE*_ function measures the difference, similar to the square Euclidean distance:

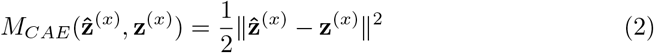

The cost function is expressed in its general form as:

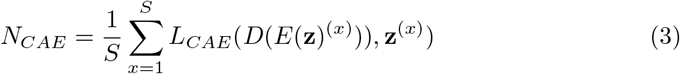

Our goal is to minimize *N*_*CAE*_ to determine optimal CAE weight parameters. In our implementation, the cost function was minimized using the Adam optimizer with a learning rate of 10^*−*3^. To avoid the dying ReLU problem [25], LeakyReLU with a negative slope of 0.2 was used to maintain gradient flow and non-linearity.

#### EfficientNet

In recent times, ENB models [15] have shown superiority over conventional DL models due to their unique architectural design and compound scaling. Their structure includes depth-wise separable convolutions, squeeze-and-excitation blocks, and inverted residual connections, allowing them to effectively capture complex features from SS (Fig. 1). Compound scaling optimizes depth, width, and resolution, resulting in accurate yet efficient models [15]. The resolution refers to the input image size, influencing the detail the model can capture. They also benefit from TL by using pre-trained weights from large datasets, allowing fast adaptation to new tasks with smaller labelled data. Their generalization ability [15] makes them suitable for image recognition tasks, including biomedical image analysis.

#### Model Training and Evaluation

The implementation involved several steps. First, the dataset was annotated and shuffled to ensure the independence of each data point and eliminate potential bias. For the CAE pipeline, the encoder output after training was used as input to XGB for classification. The data were split into training (68%), validation (12%), and testing (20%) sets. These models were trained on the training and validation sets and evaluated using the testing set with five-fold cross-validation. The model was tested individually and assessed using classification metrics such as accuracy, recall, precision, F1-score, and area under the ROC curve (AUC) scores.

Similarly, for the ENB0 pipeline, SS was used as input. We fine-tuned the pretrained ENB0 model on our dataset. This model, initially trained on large-scale datasets like ImageNet, was adapted using TL to extract features relevant to our task. We modified the final classification layer to match the two-class setting (SZ and HC), and trained the models using binary cross-entropy loss and the Adam optimizer.

### Explainable artificial intelligence

#### Gradient-weighted Class Activation Mapping (Grad-CAM)

Grad-CAM is employed in the SS-ENB0 pipeline to provide an intuitive post-hoc visualization of the model’s predictions. This technique generates a class-specific activation heatmap by using the gradient of the predicted class with respect to final convolutional features of the network, by highlighting the regions of the input that most strongly influence the output prediction [18]. Integrating Grad-CAM requires no changes to the ENB0 architecture, making it a seamless addition for explainability (Fig. 1). By indicating potentially informative EEG segments in SS, the heatmaps give experts a visual cue to validate the focus of the model, enhancing interpretability without enforcing any particular region [26].

#### SHapley Additive exPlanations (SHAP)

SHAP provides a unified approach for interpreting ML models by quantifying each feature’s contribution to the model’s predictions [19]. It uses a game-theoretic framework to assign importance values to individual features, ensuring transparency and interpretability even in complex models. In our study, SHAP was employed on the XGB classifier trained on the latent embeddings of EEG data to identify the most influential features in the classification process. This methodology, as described by Lundberg [19], enabled a detailed understanding of how features influenced the model’s decisions, providing interpretable insights into the neural signals associated with SZ.

## Results

### Models detected SZ with high accuracies and contributed brain regions

Based on the SS retrieved from the EEG data, the initial analysis includes eight ENB models, an additional six TL, and a 2D convolutional neural network (CNN) models were implemented for classification purposes. ENB0 was adopted for the later part of the analysis due to its optimal accuracy-efficiency balance. Applying Grad-CAM to ENB0 revealed delta-band power as the main driver of SZ classifications, with gamma- and theta-band activity predominating in HC predictions. Notably, the CAE has only 0.038M trainable parameters, an order of magnitude lower than ENB0-ENB07 (*≈* 4M-63M) or other TL models spanning VGG19 to Resnet50 (*≈* 2M-10M trainable parameters).

Further, after evaluating five ML classifiers (RF, XGB, SVC, KNN, and a soft-voting ensemble), XGB was selected for its combination of performance, efficiency, and interoperability. Later, SHAP was applied with five-fold cross-validation to pinpoint the EEG channels driving the CAE-XGB model. In each fold, we ranked channels by their average SHAP value on the test set and selected the 32 most and least influential features. Across all folds, the highest-impact electrodes were Cz and C3 in the central region, O1 and O2 in the occipital region, F3 in the frontal region, and Pz in the parietal region, indicating that sensorimotor, occipital, and parietal activity strongly drives SZ classification (Fig. 1). By contrast, T5 (temporal), F8 (frontal), and P4 (parietal) consistently ranked lowest (Fig. 1). These findings are consistent with known SZ-related impairments in central and parietal cortical regions [9, 27].

In our two-pronged approach, CAE-XGB achieved 95.3% accuracy and SS-ENB0 obtained 97% accuracy. Table 1 depicts the overall performance metrics for the CAE-XGB and SS-ENB0 frameworks. These results indicate that simpler models, which have fewer parameters and lower computational requirements, can perform on par with, and sometimes even surpass, more complex models. Fig. 2 illustrates interpretability and classification performance for both pipelines: SHAP values on the CAE latent space segments with XGB confusion matrix (Fig. 2A); samples of Grad-CAM overlayed on SS (top: HC, bottom: SZ) with ENBO confusion matrix (Fig. 2B).

**Table 1.** Model performance of the CAE-XGB with SS-ENB0 frameworks.

| Framework | ACC | PRE | REC | F1 | AUC |
| --- | --- | --- | --- | --- | --- |
| CAE-XGB | 0.953 | 0.971 | 0.927 | 0.949 | 0.992 |
| SS-ENB0 | 0.970 | 0.970 | 0.970 | 0.970 | 0.990 |

ACC - Accuracy, PRE - Precision, REC - Recall, F1 - F1-score, AUC - area under the receiver operating characteristic curve.

**Fig. 2.**
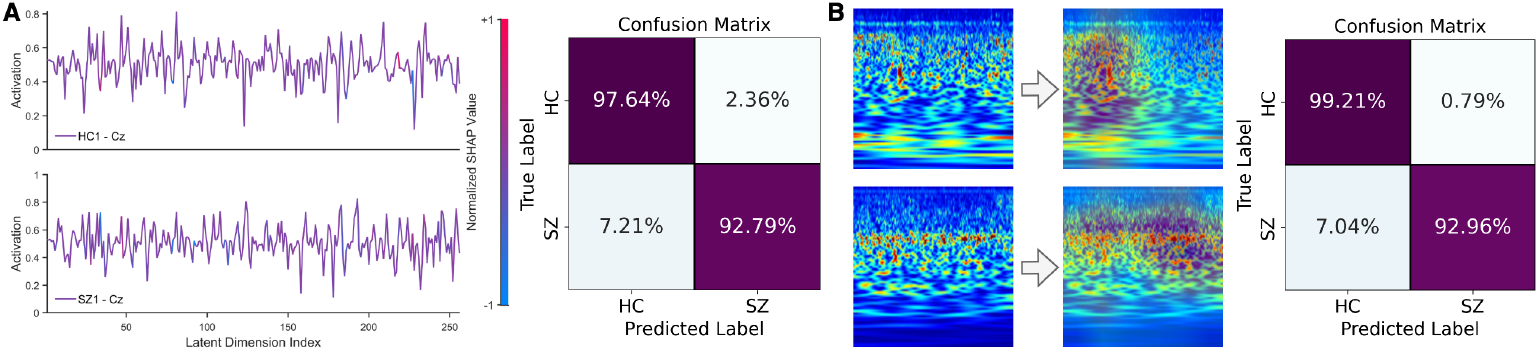
Interpretability and classification results. **A**. SHAP values projected onto the CAE latent space segments (left) and XGB confusion matrix (right). **B**. Sample Grad-CAM heatmaps (top: HC, bottom: SZ) overlaid on spectral scalograms (left) and ENB0 confusion matrix (right).

## Discussion

In this work, EEG data was used to classify SZ and evaluate the effectiveness of CAE and ENB models. The simulation findings not only offer insights into the model’s performance but also provide interpretable insights into key features which drive them. Previous studies have explored various approaches for SZ classification using EEG data. For instance, Sui et al. [28] used multi-set canonical correlation analysis with SVM and achieved 74% accuracy, while Shim et al. [29] applied a band-pass filter technique with SVM, obtaining 88.24% accuracy. Moreover, Zhang [30] employed ERP-based EEG features with RF and achieved 81.1% accuracy.

Some studies combined feature extraction techniques with DL models: Phang et al. [31] used time and frequency domain features with CNN, obtaining an accuracy of 91.69%. In another study, Siuly et al. [32] applied empirical mode decomposition with an ensemble bagged tree, achieving 89.59% accuracy. Furthermore, David et al. [33] developed a recurrent deep CNN with 89.98% accuracy, while Guo et al. [34] achieved 92% accuracy using a deep neural network. Other methods focused on integrating different feature selection and classification techniques. Akbari et al. [35] combined a two-dimensional phase space dynamic approach with KNN, obtaining an accuracy of 94.8%. Rajesh et al. [36] used symmetrically weighted local binary patterns with a logistic regression boosting classifier, achieving 91.66% accuracy.

In TL-based approaches, smoothed Pseudo-Wigner-Ville distribution combined with TL models (AlexNet and ResNet50) achieved 93.34% accuracy [37]. CNN and ResNet50, using CWT and STFT, obtained 90.64% and 79.18% accuracy, respectively. Our method demonstrated comparable performance with a wide variety of frameworks, ranging from traditional feature extraction, CNN, DL, to state-of-the-art TL-based approaches. Thus, the combination of CAE-XGB and SS-ENB pipelines not only improved classification performance but also provided insights into the model’s decision-making process, which standard DL or TL methods alone could not achieve. In comparison to these previous studies, our proposed approach not only has comparable performance in discriminating SZ from HC, but also paves the way for gaining insights into the black-box model by integrating XAI into our pipeline. While the results are promising, it is important to consider the limitations of these models. For example, ENB models require long training times and high computational power, especially in deeper variants, due to the large number of parameters. They are also sensitive to hyperparameter tuning, where factors like scaling, dropout, learning rate, and batch size must be carefully optimized. Without this, performance may degrade [15, 38]. Despite their efficiency, larger ENBs can still demand significant resources during inference, limiting real-time deployment. CAE models also have limitations. They may not capture spatial dependencies well, which are important for EEG analysis. While effective in extracting temporal features, they may underperform compared to models that better preserve inter-channel relationships. CAEs also require large and diverse datasets to learn generalizable features, and with limited or imbalanced data, performance can drop. These factors should be considered when applying such models in clinical or real-time contexts. Still, CAE-XGB and SS-ENB pipelines offer a strong balance between performance and efficiency.

To improve interpretability, SHAP was used on the CAE-XGB pipeline, which showed that channels over central (Cz, C3), occipital (O1, O2), frontal (F3), and parietal (Pz) areas were most influential, aligning with known SZ-related network changes [9, 19, 27]. Then, Grad-CAM on the ENB0 model revealed that delta-band power drove SZ predictions, while gamma and theta activity marked HC cases, complementing the spatial insights from Grad-CAM [18]. Misclassified segments often corresponded to low-frequency noise or edge artifacts, suggesting that targeted pre-processing or artifact rejection could improve robustness. Together, these insights pinpoint which scalp sites and frequency bands drive each model’s decisions, improving transparency and advancing trustworthy AI.

## Conclusion

In this work, we introduced two complementary EEG classification pipelines, employing CAE-XGB and SS-ENB0, that reduce the need for extensive pre-processing or domain-specific feature extraction of multi-channel EEG data. For the SS-ENB0 pipeline, SS from CWT were fed into ENB0; in the CAE-XGB pipeline, DWT-decomposed EEG was fed to a CAE, and its latent embeddings from the encoder served as features for classification by XGB. Our methods achieved 95.3% accuracy with CAE-XGB and 97% with SS-ENB0, demonstrating reliable, stable SZ classification.

Alongside classification performance, we addressed model interpretability by applying SHAP and Grad-CAM to provide complementary insights. SHAP analysis helped identify EEG channels most relevant to classification decisions in the CAE-XGB pipeline, while Grad-CAM revealed time-frequency regions in the SS that guided ENB0 predictions. These additions support transparency at both spatial and spectral levels, contributing to greater clinical trust in the ML framework.

## Data Availability

The source code and the generated data will be made publicly available upon publication.

## Software and Hardware Setup

The implementation used MATLAB 2021a on an Apple MacBook Pro with an M1 processor for pre-processing, and Python 3.8 with Google Colab for ML and DL model development and deployment. Source code will be made available upon publication. The original dataset [20] is openly available at (http://brain.bio.msu.ru/eegschizophrenia.htm).

## Declaration

The paper is under consideration at Pattern Recognition Letters.

